# An evaluation of the association between changes to job protection during illness leave and illness absence behaviour

**DOI:** 10.1101/2025.06.18.25329071

**Authors:** Michael Lebenbaum, Ioana Nicolau, Stuart Peacock, Jennifer Gillis, Samuel Asare

## Abstract

**Background:** Despite a growing body of research on sick leave policies, there remains a significant gap in research concerning job protections during illness leaves, which is critical in Canada since several provinces are considering or passing job-protected leave expansions. We examined three major job-protected leave expansions, Quebec (2003), Manitoba (2016), and Alberta (2018).

**Methods:** We used the Canadian Labour Force Survey data spanning from 1998 to 2022. Using a difference-in-difference approach, we examined 5-year changes in leave behaviour before and after expansions in job-protected leave in the three provinces compared to changes in provinces with less than 2 weeks of job-protected leave. We analyzed the prevalence, duration, and distribution of illness/disability absences using ordinary least squares and linear probability models.

**Results:** We found that expanding job-protected leave in Quebec was associated with decreases in the overall length of leave by 2.2 (95% CI: -3.2 to -1.5; P<0.001) weeks or 14.0% relative reduction. Similarly, expansions in Alberta and Manitoba were associated with decreases in the overall length of leave by 1.2 (95% CI: -2.1 to -0.3; P=0.016) weeks or 7.4% relative reduction. Results for absence prevalence were mixed (small increase (Quebec) (p<0.05), no significant change (Alberta-Manitoba) (p>0.05). Both expansions were associated with significant increases in absence duration consistent with the policy (i.e., 3-17-week leaves) (p<0.05).

**Conclusions:** Our results suggest that job-protected leave expansion may influence leave behaviour even in the presence of protections provided by human rights laws and without imposing large additional costs for employers or governments.

## Introduction

Employment and working conditions are important determinants of health, offering not just income but often a sense of meaning, belonging, and other intangible benefits^1^. Health and employment are intrinsically linked in a reciprocal relationship with numerous studies documenting that individuals in poor health have difficulty in maintaining employment^2,3^. Sick leave policies provide employees with the opportunity to take time off and recover before returning to work, with paid sick days being associated with reductions in the burden of unemployment, increases in return to work and lower turnover, and likely having positive health benefits for others due to a lower risk of contagion^4-6^.

While paid sick days are the most studied component of sick leave policy, these policies typically focus on wage replacement and job security for only a few days. However, many illnesses, due to their severity or the prolonged nature of their treatment, such as cancer or cardiovascular disease, can require extended illness leaves lasting weeks to several months^7^. During such extended illness leaves, income replacement is frequently available via disability or Employment Insurance programs, but job protection policies, referred to as job-protected leave (JPL), can vary significantly depending on the jurisdiction^8^. Despite extensive research on access to and effects of short-term sick pay^5^, and job protections for parental leave^9,10^, there is a lack of studies on the impacts of job protections during extended illness leave^10,11^.

Through the Employment/Labour Standards Acts (ESAs), that establish the minimum standards for employment conditions^12-21^, nearly all Canadian provinces and territories provide 1 to 26 weeks of job-protected leave, ensuring an employee’s position cannot be changed or terminated during an illness leave^8,22^. Several provinces and territories recently passed or in the process of passing legislation to extend job-protected leave to align with the December 2022 expansion of the federal Employment Insurance sickness benefit from 15 to 26 weeks^23,24^. Past changes to JPL through changes to ESA legislations, such as Quebec’s transition from 17 to 26 weeks and the adoption of 17 and 16 weeks of protection in Manitoba and Alberta respectively, occurred staggered over time. This provides an opportunity to examine the influence of illness-leave job protections on illness leave behaviour, such as absence prevalence and length. To inform on-going consultations and further expansions, we evaluated these three past extensions of job-protected leave, which occurred in 2003, 2016 and 2018 in Quebec, Manitoba and Alberta, respectively, and their impact on illness leave behaviour.

## Methods

### Data and Sample

We conducted a difference-in-difference study using repeat cross-sectional data using Labour Force Survey data using the Labour Force Survey, a monthly nationally representative survey that recruits households and gathers information primarily on labour market activities and work characteristics from a large sample (∼50,000 households, ∼80,000-120,000 individuals)^25^. The survey samples individuals aged 15 years and older living in Canada. The survey excludes persons living on reserves and other Indigenous settlements, individuals who are institutionalized or full-time members of the Canadian Armed Forces, and individuals who live in extremely remote areas, which collectively comprise less than 2% of the Canadian population^25^. While the Labour Force Survey samples individuals from all provinces and territories, the public use file used for this study was limited to individuals residing in the provinces and excludes the 3 territories, which make up ∼0.3% of the Canadian population. We restricted our sample to the years 1998-2008 to 2011-2022, due to the timing of the policy changes.

The Labour Force Survey employs a rotating panel design, selecting households into mini-panels and following the same respondents for up to six consecutive months, resulting in up to six observations per individual^25^. The survey samples individuals in all months of the year^25^. However, we relied on the public use version of the file, which is cross-sectional and does not have the information required to identify individuals over time. Therefore, we limited the sample to observations surveyed six months apart, specifically in April and October to avoid sampling multiple observations on the same individual and thus introducing serial correlation. This is the same approach as applied by other studies^9,25^.

We excluded unpaid family workers, full-time students, individuals ages 15-19 and 65+, those not employed, self-employed, workers not meeting tenure requirements of job-protected leave, unionized workers, and individuals covered by a collective agreement. Full-time students were excluded since ESAs do not apply to them and they are not eligible for most outcomes. We excluded individuals ages 15-19 and 65+ due to their low labour force participation, as well as those not employed, given the focus on illness leaves from employment. Given that ESAs do not apply to all workers, we restricted our sample to individuals who were covered or affected by ESAs, which meant excluding the self-employed, workers who did not meet tenure or eligibility requirements of job-protected leave (i.e., 90 days, which is the requirement in all three provinces), unionized workers, and individuals covered by a collective agreement. Unionized workers are typically covered by their own collective agreements that supersede the ESAs. Furthermore, excluding unionized employees addressed the lack of variables identifying the ∼5.5% of workers who are federally regulated and are covered by their own labour code with longer job-protected leave, rather than by a provincial ESA. Missing data was imputed by Statistics Canada using carry-forward, deterministic and donor imputation methods^26^.

### Primary exposure

We reviewed the ESAs of all 10 provinces to identify the timing of changes to job protections during illness leaves^12-21^. In cases where a policy change occurred mid-month, we started the policy at the beginning of the following month.

Quebec introduced 26 weeks of job-protected leave in 2003. Prior to this, Quebec provided 17 weeks of leave. In 2003, all other provinces provided less than 2 weeks of job-protected leave except for Saskatchewan, which provided 12 weeks. Manitoba introduced 17 weeks of job-protected leave in 2016, and Alberta introduced 16 weeks in 2018. All other provinces continued to provide less than 2 weeks of leave until the end of the study period in 2022.

### Outcome variables

We examined three primary outcomes to measure illness leave behaviour: any illness or disability absence, any illness or disability absence lasting three weeks or longer, and the length of illness or disability absences measured in weeks. As secondary analyses, we categorized the length of leave into segments based on the lengths of the policies passed (e.g., Quebec extended job-protected leave (JPL) from 17 to 26 weeks): 1-2 weeks, 3-11 weeks, 12-17 weeks, 18-26 weeks, and 27 or more weeks.

### Covariates

We controlled for sociodemographic characteristics, including age, sex, and education, and included survey year, survey month, and province fixed effects. Age was categorized into 5-year age intervals and education was categorized into three levels: 1) no certificate, diploma, or degree, 2) post-secondary certificate or diploma, or 3) bachelor’s degree or higher. The month variable was either April or October.

By including year fixed effects, whereby individual years are added into the model, we captured events or policy changes affecting all provinces, such as Supreme Court of Canada decisions, the Great Recession, and the COVID-19 pandemic. Controlling for the month (April/October) accounted for seasonal and weather-related differences in absences. Adding province fixed effects resulted in estimates with a within-province interpretation and helped to control for any part of the ESAs that do not change over time, as well as time-invariant human rights codes and other laws that remain unchanged.

### Analysis

Our primary analysis split the period up to 10-12 years around the time of the policy change to examine shorter-term changes after provinces passed extended job-protected leave (12+ weeks). These analyses focused on Quebec with the 26 weeks policy introduced in 2003 and Manitoba and Alberta together in the second period, given the similar levels of job-protected leave (16-17 weeks) introduced around the same time (2016/2018). For Quebec, which implemented 26-week job-protected leave in 2003, we restricted the analysis to 1998 to 2008. Comparison provinces were all other provinces, which had less than 2 weeks job-protected leave, except for Saskatchewan, which we excluded as it was the only other province with more than 2 weeks of job-protected leave. For Manitoba and Alberta, which implemented 17 and 16 weeks of job protection in 2016 and 2018 respectively, we restricted that analysis to 2011 to 2023, excluding Saskatchewan and Quebec for the same reasons.

The primary covariate of interest in this analysis was a binary variable coded as one for individuals treated with the policy in Quebec and similarly for Manitoba and Alberta. This difference-in-difference approach^27^ also included year, month and province dummies in both cases and controlled for age, sex and education.

We conducted ordinary least squares regression for length of leave and linear probability models for the rest of the binary outcomes to improve ease of interpretation and common use in Difference-in-Difference studies^27,28^. Analyses of the length of illness/disability absences included only individuals on an illness/disability absence. As a sensitivity analysis, in analyses of length of leave categories we estimated the model with multinomial logit with results shown as the average marginal effects which estimates the change in probability of each category with a change in the policy variable. All analyses applied robust standard errors clustered at the provincial level.

We also conducted descriptive analyses, examining study variables in the years pre- and post-policy implementation for intervention and comparator provinces. Additionally, we plotted trends over time in all outcomes for provinces that enacted an extensive job-protected leave during the study period (i.e., Quebec, Manitoba, Alberta) compared to all other provinces used for comparison.

All descriptive statistics were survey weighted, while regressions were not survey weighted because applying survey weights to the models may substantially increase variances unnecessarily^29^.

Ethical approval was not required as we relied on public use data modified to reduce re-identification risks.

### Results

We included a flow diagram to illustrate the change in sample size with each inclusion/exclusion step, which was prepared separately for the Quebec and Manitoba-Alberta JPL policy time windows (Appendix Figure 1). Descriptive characteristics are presented pre- and post-policy change for treatment and comparison provinces in Table 1 (Quebec) and Table 2 (Manitoba-Alberta). Following the passage of JPL, there was small increases in illness absences and an increase in absence duration categories consistent with the policy extension. When examined continuously over time, changes in the prevalence of illness/disability absences after the policy change were small and the length of illness/disability absences trended downwards following the introduction of 26-week JPL (Figure 2, Figure 3, Appendix Figure 2).

**Table 1.** Sample characteristics before and after passing of job-protected leave (JPL) for Quebec and Comparison group, Canada, 1998-2008.

|  | Quebec 5-years<br>before JPL |  | Quebec 5-years after<br>JPL |  | Comparison group<br>5-years before JPL |  | Comparison group 5-years after JPL |  |
| --- | --- | --- | --- | --- | --- | --- | --- | --- |
|  | N | % | N | % | N | % | N | % |
| Female | 20,293 | 49 | 20,088 | 49.7 | 96,114 | 48.5 | 102,386 | 48.6 |
| Age |  |  |  |  |  |  |  |  |
| 20 to 24 | 4,223 | 10.2 | 3,659 | 9.1 | 20,331 | 10.2 | 21,430 | 10.2 |
| 25 to 29 | 5,540 | 13.4 | 5,315 | 13.2 | 27,930 | 14.1 | 27,168 | 12.9 |
| 30 to 34 | 5,769 | 13.9 | 5,102 | 12.6 | 29,209 | 14.7 | 27,700 | 13.1 |
| 35 to 39 | 6,436 | 15.5 | 5,255 | 13.0 | 30,908 | 15.6 | 28,350 | 13.5 |
| 40 to 44 | 6,431 | 15.5 | 6,010 | 14.9 | 29,884 | 15.1 | 31,424 | 14.9 |
| 45 to 49 | 5,130 | 12.4 | 5,664 | 14.0 | 23,902 | 12.0 | 28,009 | 13.3 |
| 50 to 54 | 4,079 | 9.9 | 4,582 | 11.3 | 18,770 | 9.5 | 22,232 | 10.6 |
| 55 to 59 | 2,639 | 6.4 | 3,174 | 7.9 | 11,654 | 5.9 | 16,154 | 7.7 |
| 60 to 64 | 1,149 | 2.8 | 1,654 | 4.1 | 5,772 | 2.9 | 8,204 | 3.9 |
| Education |  |  |  |  |  |  |  |  |
| No post-secondary<br>certificate, diploma, degree | 17,753 | 42.9 | 15,175 | 37.5 | 89,994 | 45.4 | 88,416 | 42 |
| Post-secondary certificate<br>or diploma | 15,536 | 37.5 | 16,445 | 40.7 | 67,734 | 34.1 | 73,081 | 34.7 |
| University degree or higher | 8,107 | 19.6 | 8,794 | 21.8 | 40,633 | 20.5 | 49,174 | 23.3 |
| Province |  |  |  |  |  |  |  |  |
| Newfoundland |  |  |  |  | 3,198 | 1.6 | 3,370 | 1.6 |
| Prince Edward Island |  |  |  |  | 1,001 | 0.5 | 1,089 | 0.5 |
| Nova Scotia |  |  |  |  | 7,617 | 3.8 | 7,930 | 3.8 |
| New Brunswick |  |  |  |  | 6,320 | 3.2 | 6,412 | 3 |
| Québec | 41,396 | 100 | 40,414 | 100 | - | - | - | - |
| Ontario |  |  |  |  | 110,648 | 55.8 | 115,932 | 55 |
| Manitoba |  |  |  |  | 8,736 | 4.4 | 8,662 | 4.1 |
| Alberta |  |  |  |  | 29,838 | 15.0 | 33,615 | 16.0 |
| British Columbia |  |  |  |  | 31,002 | 15.6 | 33,660 | 16.0 |
| Survey Year |  |  |  |  |  |  |  |  |
| 1998 | 3,912 | 9.4 |  |  | 18,287 | 9.2 |  |  |
| 1999 | 8,004 | 19.3 |  |  | 37,985 | 19.1 |  |  |
| 2000 | 8,191 | 19.8 |  |  | 39,348 | 19.8 |  |  |
| 2001 | 8,380 | 20.2 |  |  | 40,374 | 20.4 |  |  |
| 2002 | 8,628 | 20.8 |  |  | 41,197 | 20.8 |  |  |
| 2003 | 4,281 | 10.3 | 3,854 | 9.5 | 21,169 | 10.7 | 20,186 | 9.6 |
| 2004 |  |  | 8,179 | 20.2 |  |  | 41,220 | 19.6 |
| 2005 |  |  | 8,086 | 20.0 |  |  | 41,405 | 19.7 |
| 2006 |  |  | 8,013 | 19.8 |  |  | 42,658 | 20.2 |
| 2007 |  |  | 8,183 | 20.2 |  |  | 43,208 | 20.5 |
| 2008 |  |  | 4,100 | 10.1 |  |  | 21,994 | 10.4 |
| Survey Month |  |  |  |  |  |  |  |  |
| April | 20,721 | 50.1 | 20,098 | 49.7 | 100,202 | 50.5 | 105,662 | 50.2 |
| October | 20,675 | 49.9 | 20,316 | 50.3 | 98,159 | 49.5 | 105,009 | 49.8 |
| Any illness absence | 804 | 1.9 | 859 | 2.1 | 2,672 | 1.3 | 2,991 | 1.4 |
| 3+ week illness absence | 617 | 1.5 | 665 | 1.6 | 1,880 | 0.9 | 2,059 | 1.0 |
| Weeks away among workers on illness/disability absence (Mean, SE) | 18.97 | 1.15 | 17.05 | 0.90 | 14.92 | 0.51 | 15.40 | 0.48 |
| Weeks away (among workers on illness/disability absence) |  |  |  |  |  |  |  |  |
| 1-2 weeks | 199 | 23.3 | 219 | 22.5 | 881 | 29.7 | 1,078 | 31.2 |
| 3-11 weeks | 312 | 36.4 | 365 | 37.5 | 1,093 | 36.8 | 1,180 | 34.1 |
| 12-17 weeks | 81 | 9.5 | 109 | 11.2 | 295 | 9.9 | 334 | 9.7 |
| 18-26 weeks | 78 | 9.2 | 80 | 8.2 | 207 | 7.0 | 282 | 8.1 |
| 27+ weeks | 186 | 21.7 | 200 | 20.6 | 494 | 16.6 | 587 | 17 |

Legend: JPL = Job protected leave

**Table 2.**
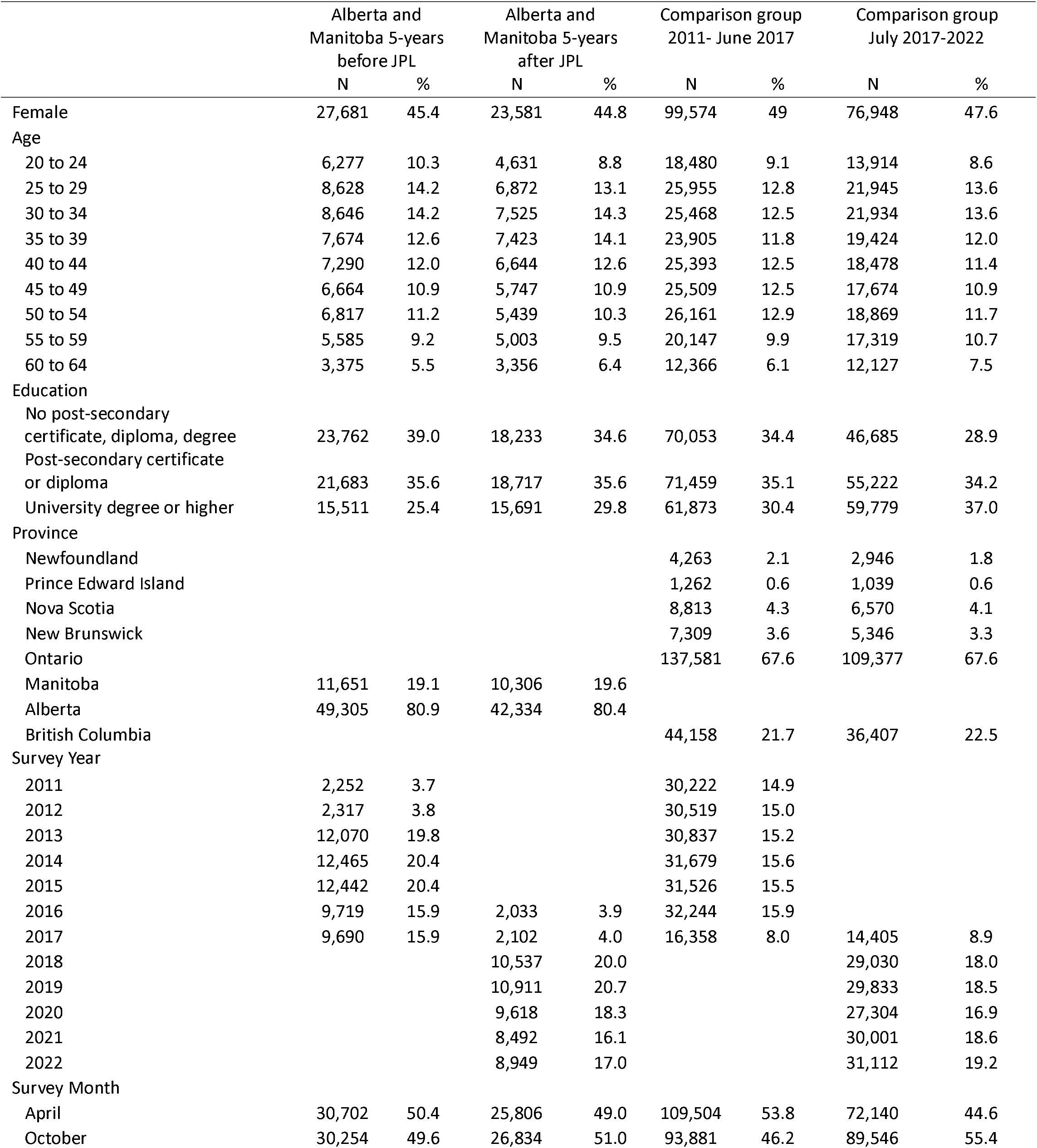
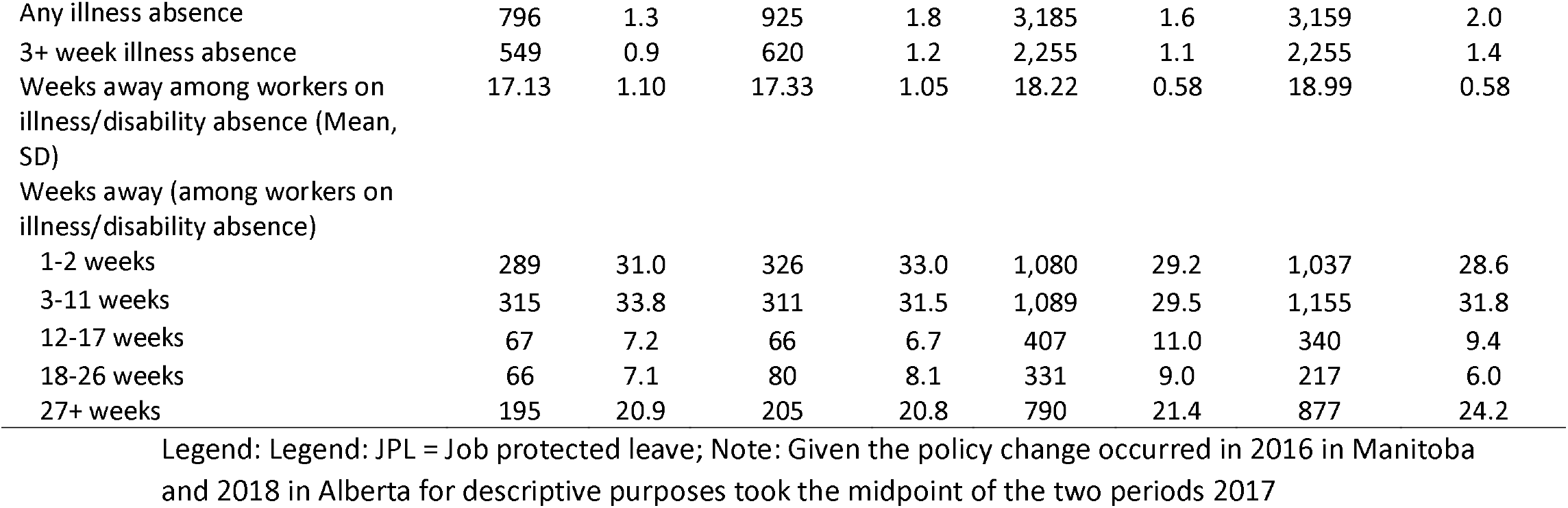
Sample characteristics before and after passing of job-protected leave (JPL) for Alberta and Manitoba combined and comparison group, Canada, 2011-2022.

**Figure 1.**
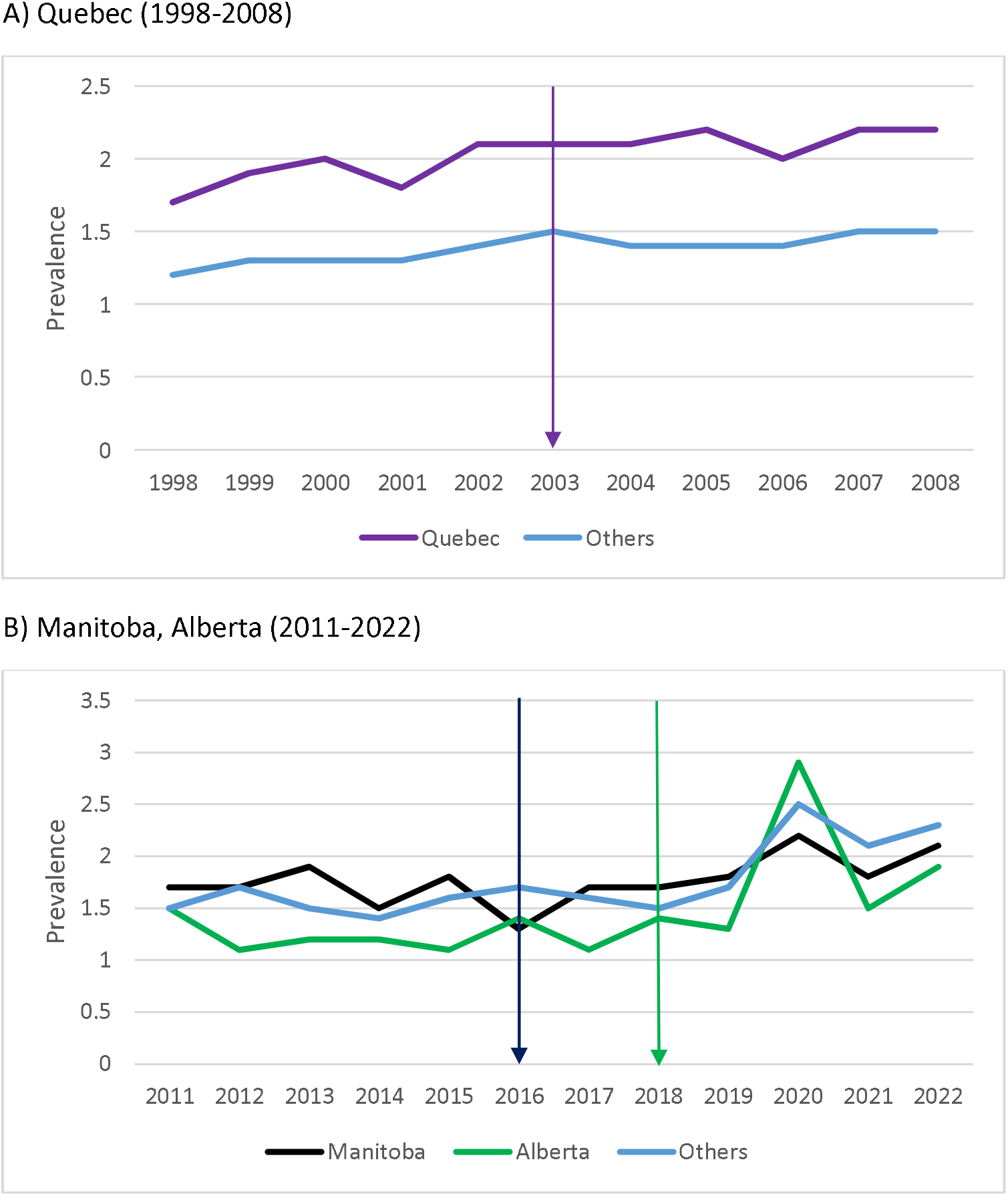
Trends over time in the prevalence of absences due to illness/disability for A) Quebec vs. other provinces and B) Manitoba and Alberta vs. other provinces Legend: Each line highlights the corresponding policy change with the arrow colour matching the colour of the province/line. Others include all other provinces excluding Saskatchewan for A) Quebec and Saskatchewan and Quebec for B) Manitoba and Alberta.

**Figure 2.**
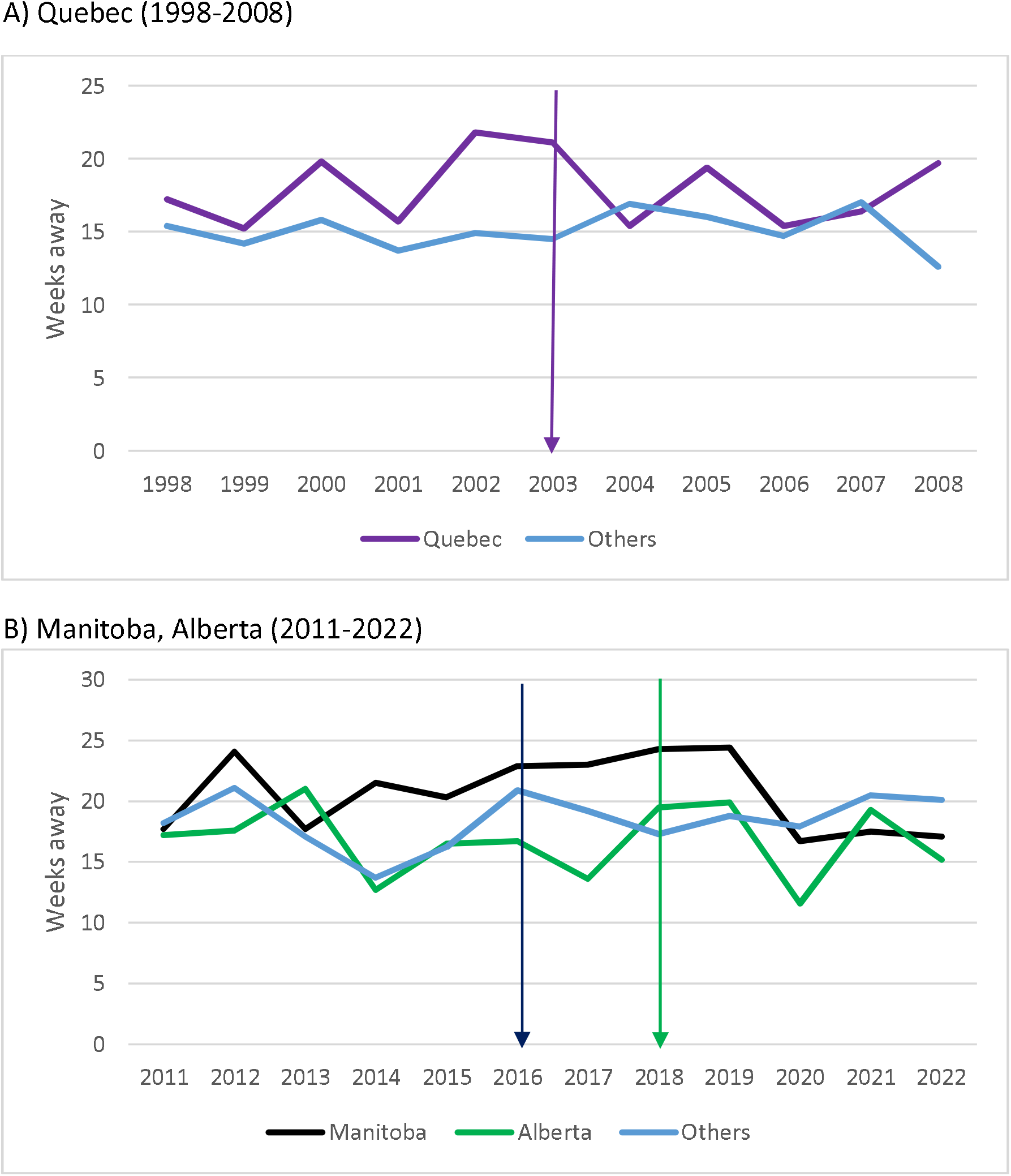
Trends over time in weeks away among illness absence A) Quebec vs. other provinces and B) Manitoba and Alberta vs. other provinces Legend: Each line highlights the corresponding policy change with the arrow colour matching the colour of the province/line. Others include all other provinces excluding Saskatchewan for A) Quebec and Saskatchewan and Quebec for B) Manitoba and Alberta.

We found that Quebec’s 26-week JPL policy was associated with the prevalence of illness/disability absences (β=0.002, p=0.047; 13.3% increase) and 3+ week illness/disability (β=0.002, p=0.008; 18.2% increase) (Table 3). We did not find any significant associations between Alberta and Manitoba’s 16–17-week JPL policy and illness/disability absence prevalence.

**Table 3.** Primary analysis of job protected leave (JPL) policy changes examining changes in illness/disability prevalence, length of leave and distribution of leave.

| *Outcome* | *JPL Policy Change* | $\beta$ | 95% CI | P-Value |
| --- | --- | --- | --- | --- |
| Illness/disability | 16-17 weeks | -0.001 | -0.004,0.003 | 0.661 |
| Absence | 26 weeks | 0.002 | -0.000,0.004 | 0.047 |
|  | Mean 16-17 week period | 0.017 |  |  |
|  | Mean 26 week period | 0.015 |  |  |
| 3+ week | 16-17 weeks | -0.001 | -0.004,0.002 | 0.397 |
| illness/disability | 26 weeks | 0.002 | 0.001,0.003 | 0.007 |
| absence | Mean 16-17 week period | 0.012 |  |  |
|  | Mean 26 week period | 0.011 |  |  |
| Overall Length of | 16-17 weeks | -1.178 | -2.054,-0.302 | 0.016 |
| leave | 26 weeks | -2.230 | -3.153,-1.450 | <0.001 |
|  | Mean 16-17 week period | 18.37 |  |  |
|  | Mean 26 week period | 15.96 |  |  |
| *Length of leave divided into categories* |  |  |  |  |
| 1-2 week leaves | 16-17 weeks | 0.026 | -0.026,0.078 | 0.271 |
|  | 26 weeks | -0.011 | -0.035,0.012 | 0.301 |
|  | Mean 16-17 week period | 0.296 |  |  |
|  | Mean 26 week period | 0.283 |  |  |
| 3-11 week leaves | 16-17 weeks | -0.054 | -0.113,0.006 | 0.070 |
|  | 26 weeks | 0.047 | 0.029,0.066 | <0.001 |
|  | Mean 16-17 week period | 0.310 |  |  |
|  | Mean 26 week period | 0.358 |  |  |
| 12-17 week leaves | 16-17 weeks | 0.027 | 0.008,0.047 | 0.012 |
|  | 26 weeks | -0.002 | -0.015,0.010 | 0.702 |
|  | Mean 16-17 week period | 0.095 |  |  |
|  | Mean 26 week period | 0.100 |  |  |
| 18-26 week leaves | 16-17 weeks | 0.023 | -0.001,0.046 | 0.056 |
|  | 26 weeks | -0.008 | -0.033,0.018 | 0.512 |
|  | Mean 16-17 week period | 0.075 |  |  |
|  | Mean 26 week period | 0.079 |  |  |
| 27+ week leaves | 16-17 weeks | -0.023 | -0.072,0.026 | 0.304 |
|  | 26 weeks | -0.026 | -0.040,-0.012 | 0.002 |
|  | Mean 16-17 week period | 0.224 |  |  |
|  | Mean 26 week period | 0.180 |  |  |

Legend: JPL = job-protected leave; Analyses of length of leave and each of the length of leave categories were conducted among individuals who are on an illness/disability absences. Models were conducted with ordinary least squares regression (length of leave) or linear probability models (all other models). Mean refers to mean level of the dependent variable. The 16-17 week (2011-2023) and 26 week (1998-2008) JPLs are estimated in separate models across different time periods. Models control for age, sex, education, year, month and province. In analyses of 26 week policy change, all other provinces except Saskatchewan are included in the reference group, while in analysis of 16-17 week policy, all other provinces except Quebec and Saskatchewan are included in the reference group.

We found an overall 14.0% decrease in the length of leaves after the passage of the 26-week JPL policy (β=-2.230, p<0.001) (Table 3). This decrease was likely driven by a 14.4% decrease in 27+ week leaves (β=-0.026, p=0.002). After the passage of the 16-17 week JPL policy, we found a smaller significant reduction in length of leave (β=-1.178, p=0.016). Additionally, we found the 26-week policy was significantly associated with 3-11-week leaves (β= 0.047, p<0.001) and 16-17 week policies were significantly associated with 12-17 week leaves (β= 0.027, p=0.012). Analyses of length of leave categories conducted with multinomial logit provided largely the same conclusion with the exception that the 16-17 week policy was now significantly negatively associated with 3-11 week leaves and positively associated with 18-26 week leaves (p<0.05) (Appendix Table 1).

### Interpretation

### Summary of findings

We observed that the implementation of 26 weeks of job-protected leave was associated with a significant reduction in the length of JPLs, likely driven by a reduction of lengthy (27+ week) illness/disability absences. Generally, we found that extended job-protected leave (16+ weeks) was associated with increases in parts of the distribution of illness/disability absence length that were aligned with JPL policies enacted. We found mixed findings with respect to JPL and absence prevalence.

Our findings have several key implications. We observed an increase in illness/disability absences of less than 27 weeks, combined with a reduction in absences longer than 26 weeks. Based on this finding, we hypothesize that the 26-week JPL policy may have facilitated quicker recovery from illness. By encouraging leaves consistent with the JPL policies, individuals may have been able to recover more effectively, thereby reducing the need for extended absences beyond 26 weeks. Similarly, cuts to paid sick leave have been associated with increases in sick leave duration^30^, potentially due to sick leaves role in promoting access to health care, which can facilitate recovery^31^. Furthermore, these findings also indicate that changes in the provincial ESAs may influence illness/disability absences, despite the existence of provincial human rights codes or acts, which also provide protections during illness leaves, where terminations would be considered discrimination on the grounds of disability. This is important given that for human rights codes, individuals would need to enforce their own rights, which may be challenging for someone experiencing the onset of a severe illness. Lastly, given the generally insignificant or small increase in the prevalence of illness/disability absences along with decreases in leave length, our results suggest that the costs to businesses and governments of implementing 16-17 and 26 weeks of JPL are unlikely to be large and there may be some cost offsets. Although an important finding, employment insurance coverage only lasted 15 weeks during the study period, after which leaves would be unpaid for those without disability insurance. Given that Employment Insurance sickness benefits were extended to 26 weeks in 2022, studies should evaluate the impact of recent and future extensions to job-protected leave.

### Comparison to prior studies

Given a lack of studies specifically addressing job protections during longer-term illness leave, we focused on evidence related to other aspects of illness leave. A key component of sick leave policy is the provision of paid sick days, which is the most extensively studied aspect of sick leave policy. With the gradual introduction of paid sick days across the United States, a growing body of research has highlighted effects on, or associations with, absenteeism, presenteeism, access to care, and health. Studies generally showed increases in absenteeism^5^; however, they also typically found positive associations with other employment, business, health, and health care utilization outcomes^5,31,32^.

Income replacement rates are another commonly studied aspect of sick leave policies. Several European studies, including those from Italy, Sweden and other countries, have examined the impact of policies altering the income replacement rates of sickness leaves^33,34^. In Europe, replacement rates tend to be more generous relative to those in Canada. These studies generally found that more generous replacement rates lead to increased absenteeism, while less generous rates resulted in decreased absenteeism^33,34^.

Although there has been significant research into the effects of paid sick days and replacement rates during longer illness leaves, there is a notable lack of studies examining the effects of changes in job protections during longer-term illness leaves. Our study is the first to investigate policies focused solely on introducing additional protections^11^. Our findings are generally consistent with evaluations of paid sick days, suggesting that both policies may lead to increased absences but do not appear to incur substantial costs to businesses or governments.

## Limitations

This manuscript is not without limitations. First, we did not have access to administrative data on medical leave, so we relied on self-reports of illness or disability absences, which were consistently measured for all provinces throughout our study. Previous studies have shown mixed results on the validity of self-reported illness absences compared to administrative records, but findings suggest we may have missed some illness/absences and that recall bias may have affected reports of longer leaves^35^. However, we would not expect this bias to have changed over time, or have differed between intervention and comparison provinces. Additionally, other recent studies have relied on similar self-reported sickness absence data as a measure of sick leave^36,37^. Second, the data only included information on any illness/disability, preventing us from examining illness absences by specific conditions. Future research should investigate the effects of changes on job-protected leave on the leave behaviour across specific chronic conditions. Third, provinces made other corresponding leave policy changes, such as the simultaneous introduction of 10 days of family leave in Quebec in 2003 and critical illness of children leave in Alberta in 2018, which could potentially influence absences. However, by focusing our study on illness/disability absences rather than all absences, we likely reduced bias due to these simultaneous policy changes. Fourth, we lacked data on awareness of the policy changes. Lastly, we found some evidence against parallel trends for certain outcomes, which may introduce some bias into our estimates.

## Conclusions

In conclusion, the introduction of 16+ weeks of leave was associated with a reduction in the length of absences and was linked to changes to illness absence lengths that align with the intended policy outcomes. There was mixed findings with respect to absence prevalence. Our results suggest that adjustments to job-protected leave may influence illness absences without appearing to incur substantial costs to governments or businesses.

## Supporting information

All appendicies

## Data Availability

The study used ONLY openly available human data that were accessed via email to Statistics Canada and originally located at: https://www150.statcan.gc.ca/n1/en/catalogue/71M0001X

