## Supplementary material for "An evaluation of the association between changes to job protection during illness leave and illness absence behaviour": All appendicies

Appendix Figure 1A/B. Study flow diagram of inclusion/exclusions for Quebec and Manitoba-Alberta study policy windows

A) Quebec Policy Period (1998-2008)

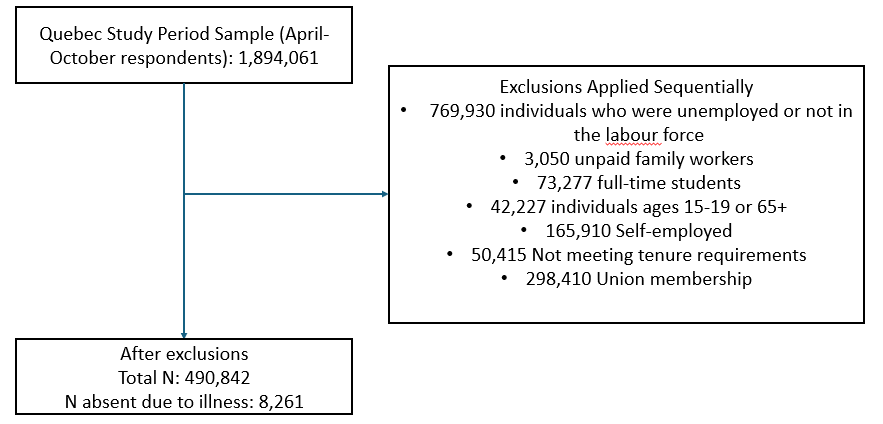

B) Manitoba-Alberta Policy Period (2011-2022)

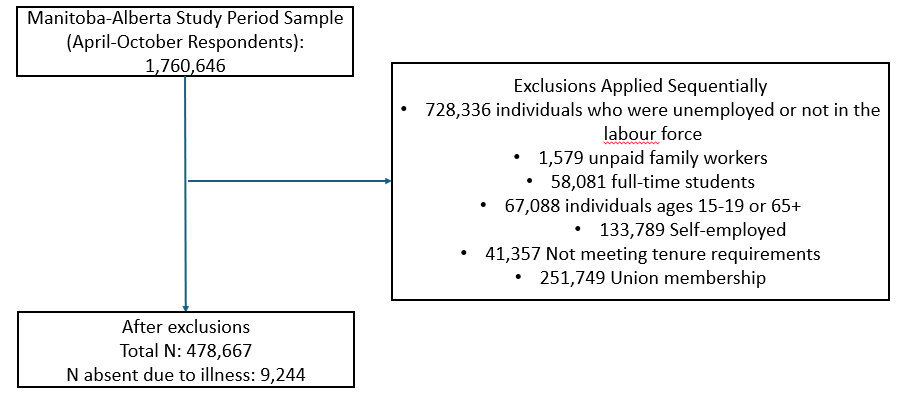
Note: Full sample already excludes Saskatchewan in case of Quebec period and Saskatchewan and Quebec in case of Alberta-Manitoba period. Quebec periods is from 1998-2008 while Manitoba-Alberta period is from 2011-2022.

Appendix Figure 2. Trends over time in 3+ week illness absence for A) Quebe vs. other provinces and B) Manitoba and Alberta vs. other provinces

A) Quebec (1998-2008)

|  | Quebec | Others |
| --- | --- | --- |
| 1998 | 1.2 | 0.9 |
| 1999 | 1.4 | 0.9 |
| 2000 | 1.6 | 1 |
| 2001 | 1.3 | 0.9 |
| 2002 | 1.7 | 1 |
| 2003 | 1.8 | 1 |
| 2004 | 1.7 | 1 |
| 2005 | 1.7 | 1 |
| 2006 | 1.5 | 0.9 |
| 2007 | 1.6 | 1.1 |
| 2008 | 1.6 | 1 |

B) Manitoba, Alberta (2011-2022)

|  | Manitoba | Alberta | Others |
| --- | --- | --- | --- |
| 2011 | 1.1 | 0.9 | 1 |
| 2012 | 1.2 | 0.8 | 1.3 |
| 2013 | 1.5 | 0.8 | 1.1 |
| 2014 | 1.1 | 0.8 | 0.9 |
| 2015 | 1.2 | 0.7 | 1 |
| 2016 | 0.8 | 1.1 | 1.3 |
| 2017 | 1.4 | 0.7 | 1.2 |
| 2018 | 1.2 | 1 | 1.2 |
| 2019 | 1.3 | 0.9 | 1.2 |
| 2020 | 1.6 | 1.8 | 2 |
| 2021 | 1 | 1.1 | 1.4 |
| 2022 | 1.3 | 1.1 | 1.4 |

Legend: Each line highlights the corresponding policy change with the arrow colour matching the colour of the province/line. Others include all other provinces excluding Saskatchewan for A) Quebec and Saskatchewan and Quebec for B) Manitoba and Alberta.

Appendix Table 1. Primary analysis of job-protected leave (JPL) policy changes examining changes in the distribution of leave, estimated with multinomial logit

| *Outcome* | *JPL Policy Change* | β | 95% CI | P-Value |
| --- | --- | --- | --- | --- |
| *Length of leave divided into categories* | | |  |  |
| 1–2-week leaves | 16-17 weeks | 0.023 | -0.023,0.070 | 0.330 |
|  | 26 weeks | -0.012 | -0.030,0.006 | 0.202 |
|  | Mean 16–17-week period | 0.296 |  |  |
|  | Mean 26-week period | 0.283 |  |  |
| 3–11-week leaves | 16-17 weeks | -0.054 | -0.095,-0.012 | 0.011 |
|  | 26 weeks | 0.047 | 0.030,0.063 | <0.001 |
|  | Mean 16–17-week period | 0.310 |  |  |
|  | Mean 26-week period | 0.358 |  |  |
| 12–17-week leaves | 16-17 weeks | 0.030 | 0.006,0.054 | 0.015 |
|  | 26 weeks | -0.003 | -0.014,0.007 | 0.547 |
|  | Mean 16–17-week period | 0.095 |  |  |
|  | Mean 26-week period | 0.100 |  |  |
| 18–26-week leaves | 16-17 weeks | 0.025 | 0.002,0.048 | 0.036 |
|  | 26 weeks | -0.007 | -0.028,0.015 | 0.541 |
|  | Mean 16–17-week period | 0.075 |  |  |
|  | Mean 26-week period | 0.079 |  |  |
| 27+ week leaves | 16-17 weeks | -0.024 | -0.065,0.017 | 0.260 |
|  | 26 weeks | -0.025 | -0.036,-0.014 | <0.001 |
|  | Mean 16–17-week period | 0.224 |  |  |
|  | Mean 26-week period | 0.180 |  |  |

Legend: JPL = job-protected leave; Analyses of each of the length of leave categories were conducted among individuals who are on an illness/disability absences. Models were conducted with multinomial logit. Mean refers to mean level of the dependent variable. The 16-17 week (2011-2023) and 26 week (1998-2008) JPLs are estimated in separate models across different time periods. Models control for age, sex, education, year, month and province. In analyses of 26 week policy change, all other provinces except Saskatchewan are included in the reference group, while in analysis of 16-17 week policy, all other provinces except Quebec and Saskatchewan are included in the reference group. Results are shown as average marginal effects which estimates the change in probability of each category with a change in the policy variable.
